# Persistent lymphocytopenia does not increase nosocomial infection risk in the ICU

**DOI:** 10.1101/2020.07.14.20153601

**Authors:** Meri R.J. Varkila, Louise Marrec, Thomas Daix, Imo E Hoefer, Saskia Haitjema, Marc J.M. Bonten, Olaf L. Cremer

**Author notes:** **Corresponding author:** Meri R.J. Varkila, UMC Utrecht, Huispostnummer F06.149, Heidelberglaan 100, 3584 CX Utrecht, The Netherlands.

## Abstract

**Background:** Lymphocytopenia is frequent in critically ill patients and has been associated with an increased risk of nosocomial infections and death in the ICU. Immunotherapies to promote recovery of lymphocyte counts have therefore been proposed. However, it is unknown if lymphocytopenia is a direct cause of ICU-acquired infections and death, or merely a marker of disease severity. We set out to study the prevalence, temporal evolution, and clinical correlates of lymphocytopenia in ICU patients, and estimate the attributable risk of lymphocytopenia in ICU-acquired infections.

**Methods:** We assessed the association between persistent lymphocytopenia (absolute lymphocyte counts <1×10^9/L on day 4) and ICU-acquired infections using multivariable competing risk Cox-regression analyses.

**Results:** Among 2302 patients admitted to a Dutch tertiary ICU having sepsis, trauma, or major surgery between 2011 and 2018, persistent lymphocytopenia was observed in 980 (42.6%) subjects. Lymphocyte counts remained relatively stable during early ICU admission, and the median duration of lymphocytopenia was 3 (IQR 1-6) days among exposed patients. ICU-acquired infections occurred in 239 (18.1%) patients without and 214 (21.8%) patients with persistent lymphocytopenia (p=0.03). However, in multivariable survival analysis persistent lymphocytopenia was not associated with infection occurrence, either directly (adjusted cause-specific HR 1.08, 95% CI, 0.90–1.31) or indirectly (subdistribution HR 1.09, 95% CI, 0.91–1.32). Sensitivity analyses did not alter these findings.

**Conclusion:** Persistent lymphocytopenia was not associated with a higher incidence rate of nosocomial infections in critically ill patients. This challenges the rationale for using absolute lymphocyte counts as a therapeutic target to prevent ICU-acquired infections.

## Introduction

Critical illness frequently results from infectious or sterile insults that are accompanied by a systemic immune response, causing life-threatening organ dysfunction. In addition to an excessive pro-inflammatory reaction, this response is characterized by concomitant activation of anti-inflammatory pathways [1]. When these counterbalancing responses become dysregulated, quantitative and qualitative alterations in innate and adaptive immune cell populations may occur that render the host susceptible to nosocomial infections [2-4], causing subsequent morbidity and mortality in the ICU [5].

A hallmark of critical illness-induced immunosuppression is anergy and apoptosis of lymphocytes [6-11]. Lymphocytopenia has been implicated in the development of infections through a combination of diminished T-cell and B-cell responses, including an impaired ability to generate immunological memory against invading microorganisms, support the production of specific antibodies, and promote microbicidal activity to clear the pathogen, as well as recognize and kill infected cells [12-14]. While most of these insights are based on *in vitro* studies, animal experiments, and postmortem histopathological observations [15], several clinical epidemiological studies have also reported associations between (both quantitative and functional) deficits in lymphocyte populations and an increased risk of intensive care unit-acquired infections (ICU-AI) and/or mortality in critically ill patients [3-4, 11, 16-21]. However, most of these studies suffered from methodological shortcomings and lack of statistical power that limited their ability to appropriately adjust for confounders and competing events, such as death or discharge (which may prevent the occurrence of new-onset infection in the ICU). It therefore remains unclear whether lymphocytopenia is a marker of disease severity or actually constitutes a cause of new or recurrent infections in ICU patients. Moreover, most prior studies have focused on specific subpopulations of critically ill patients and lacked repeated measurement of lymphocyte counts [4, 16-20]. As a result, comprehensive knowledge on the incidence, temporal evolution, and clinical correlates of lymphocytopenia in a general ICU population is lacking. Evidence supporting the role of lymphocytopenia as an independent risk factor for new-onset secondary infections in the ICU thus remains speculative.

## Methods

### Study design and population

This study was conducted within the framework of the Molecular Diagnosis and Risk Stratification of Sepsis (MARS) cohort, for which the institutional review board approved an opt-out method of informed consent (protocol number 10-056C). For the current analysis, we included critically ill adults (>18 years) who had been admitted to the University Medical Center Utrecht (UMCU, a tertiary care hospital in the Netherlands) between January 2011 and December 2018, having an ICU admission related to trauma, major surgery or sepsis with a length of stay greater than 4 days. Exclusion criteria were known prior immunodeficiency (defined as a history of solid organ or stem cell transplantation, seropositivity for the human immunodeficiency virus, hematological malignancy, functional asplenia or recent splenectomy, and any known humoral or cellular immune deficiency), pregnancy, and ICU-admission following cardiac surgery or burn injuries.

### Exposure

Routine hematological analysis was performed on a daily basis while patients were in the ICU. White blood cell (WBC) classification and enumeration was based on morphological characteristics that are routinely measured for each sample using a Cell-Dyn Sapphire hematology analyzer (Abbott Diagnostics, Santa Clara, CA, USA). Data were extracted from the Utrecht Patient Oriented Database (UPOD) as described elsewhere [22]. Absolute lymphocyte counts (ALC) were categorized into three predefined classes: normal (≥1.0 ×10^9/L), moderate lymphocytopenia (0.5—1.0 ×10^9/L), and severe lymphocytopenia (<0.5 ×10^9/L) [4, 16, 23]. If multiple counts were measured on a single day, the mean value for that day was used. However, classification of lymphocytopenia upon ICU admission was based on the first available ALC only (i.e., measured within 24 hours). Persistent lymphocytopenia was defined as ALC <1.0 ×10^9/L present on day 4.

### Outcome

The primary outcome of the study was the first occurrence of newly-acquired infection with onset later than 96 hours after ICU admission. To this end, (presumed) infectious events were prospectively registered upon each occasion that antimicrobial therapy was initiated in the ICU. For all episodes, the *post-hoc* likelihood of true infection was subsequently adjudicated by dedicated MARS observers, using available post-hoc clinical, radiological and microbiological information and applying criteria derived from the Center of Disease Control and International Sepsis Forum Consensus Conference [24]. For the primary outcome, all events having a post-hoc plausibility of infection rated at least possible were considered (i.e., only events with plausibility ‘none’ were ruled out). Since viral reactivation is not routinely monitored in our ICU, episodes of viral reactivation were not included as outcomes. As a secondary study outcome, we analyzed ICU mortality.

### Data analysis

To investigate whether day-4 ALC adequately represented prolonged exposure (i.e., persistence of lymphocytopenia), we visualized the temporal evolution of lymphocyte counts during the first 4 days in ICU using alluvial diagrams. Furthermore, we assessed the relation between various patient and/or disease characteristics and the occurrence of lymphocytopenia using descriptive statistics (i.e., χ2 and Wilcoxon rank-sum test, as appropriate).

To estimate the association between persistent lymphocytopenia and the incidence of ICU-AI we used Cox proportional hazard models, for which ICU death and discharge were considered competing events. A competing-risk survival analysis provides two measures of association: the cause-specific hazard ratio (CSHR), which estimates the direct effect of lymphocytopenia on each of the modeled outcomes, and the subdistribution hazard ratio (SHR), which measures for each point in time the cumulative risk for the event of interest among patients who did not have an event before that time.

Baseline variables that showed potentially relevant differences between exposed and non-exposed groups (i.e., *p* value <0.30) were evaluated as a potential confounders for this analysis. However, as no single variable met statistical criteria to suggest actual confounding, we performed the final multivariable adjusted analysis using only covariates derived from literature [4, 16, 25]. These included age, sex, Charlson comorbidity index, acute physiology and chronic health evaluation (APACHE) IV score, SOFA score on day 4, chronic use of immunosuppressive medications (prednisone >0.1 mg/kg for >3 months, prednisone >75 mg/day for >1 week, or equivalent), exposure to chemo- or radiotherapy in the year before ICU admission, and high-dose corticosteroid therapy during the first four days in ICU. Since any antimicrobial therapy initiated before day-4 could possibly delay (or mask) a new infectious event in the ICU, we also included this as a possible confounder in the model.

Several sensitivity analyses were performed to test the robustness of our primary findings. First, we used different definitions for persistent lymphocytopenia (i.e., by varying the ALC thresholds and required duration of exposure). Second, to take both severity and duration of lymphocytopenia into account simultaneously, we constructed a ‘lymphocytopenia score’ that assigned daily points to patients according to their level of lymphocytopenia during the first 4 days in ICU (i.e., 0 for normal ALC, 1 for moderate lymphocytopenia, and 2 for severe lymphocytopenia). We then used this score as an alternative measure of exposure in a sensitivity analysis. Third, to investigate whether directional trends in lymphocyte count might impact ICU-AI risk, we used the day-4 ALC/first ALC ratio (which reflects the slope of temporal change of ALC). Finally, to test for possible effect modification by admission diagnosis we performed stratified analyses among various subgroups of patients.

Daily ALC values were missing on 93 occasions during the first four days in ICU. These data were assumed to be missing at random and were imputed using a last observation carried forward method. Analyses were performed using SAS 9.4 (SAS Institute, Cary, NC, USA) and R software (R Foundation for Statistical Computing, Vienna, Austria).

## Results

During the study period a total of 11075 University Medical Center Utrecht patients were enrolled in the MARS cohort, 2302 of whom met the inclusion criteria for the current analysis (figure 1). Among these, the main reason for ICU admission was sepsis, trauma, and major surgery in 1123 (48.8%), 423 (18.4%) and 1206 (52.4%) cases, respectively.

**Figure 1.**
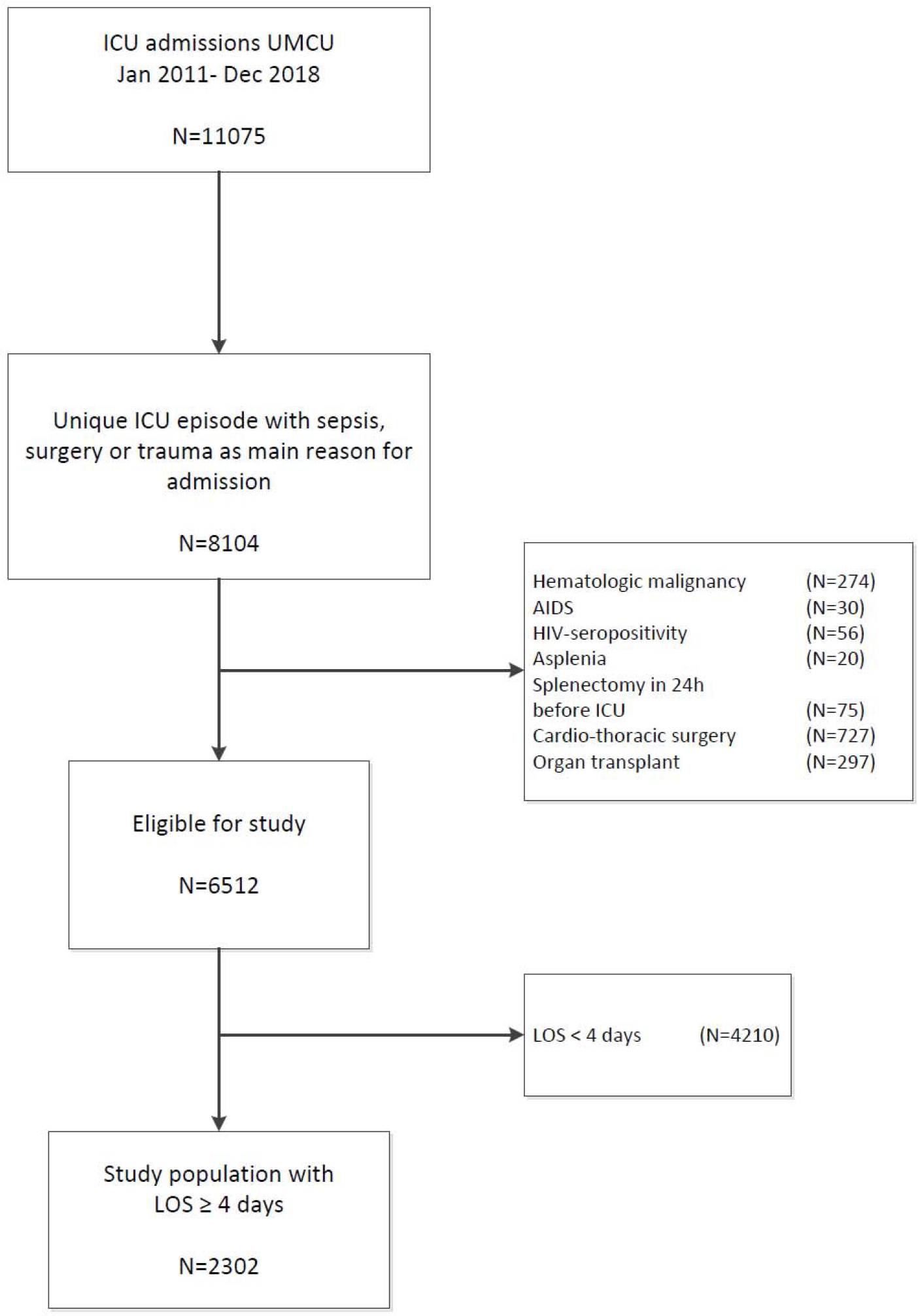
Flowchart of patient inclusion.

Persistent lymphocytopenia (ALC <1.0 ×10^9/L on day 4) was observed in 980 (42.6%) patients. Baseline characteristics of the study population are shown in table 1, stratified according to presence of persistent lymphocytopenia. Patients with persistent lymphocytopenia were older, more often male, had more chronic comorbidities, higher APACHE IV and SOFA scores, and more frequently suffered from septic shock. In addition, a larger proportion were treated with high-dose corticosteroids during the first days in ICU, or had been exposed to prior immunosuppressive therapy. Lymphocytopenia was classified as moderate (ALC 0.5-1.0 ×10^9/L) and severe (ALC <0.5 ×10^9/L) in 771 (33.5%) and 209 (9.1%) patients, respectively.

**Table 1.** Characteristics of patients by absolute lymphocyte count

| **Table 1** Characteristics of patients by absolute lymphocyte count |  |  |  |
| --- | --- | --- | --- |
| Demographics and Characteristics of ICU stay | Normal ALC<br>(ALC $\geq 1 \times 10^9/L$)<br>N=1322 | Persistent lymphocytopenia<br>(ALC $< 1 \times 10^9/L$)<br>N=980 | p |
| Age, median (IQR) | 60 (46-70) | 65 (54-73) | <0.001 |
| Male, N (%) | 810 (61.3) | 702 (71.6) | <0.001 |
| **Medical history** |  |  |  |
| Congestive Heart Failure | 160 (12.1) | 123 (12.6) | 0.80 |
| Renal Insufficiency | 82 (6.2) | 114 (11.6) | <0.001 |
| Malignancy | 194 (14.7) | 253 (25.8) | <0.001 |
| Immunosuppressant therapy prior to ICU admission<sup>a</sup> | 106 (8.0) | 177 (18.1) | <0.001 |
| Charlson Comorbidity Index | 1 (0-2.0) | 2 (0-3.0) | <0.001 |
| **ICU admission characteristics** |  |  |  |
| Sepsis at admission | 657 (49.7) | 466 (47.5) | 0.33 |
| Trauma | 236 (17.9) | 187 (19.1) | 0.49 |
| Post-operative admission | 679 (51.4) | 527 (53.8) | 0.27 |
| APACHE IV score | 65 (50-83) | 68 (53-87) | 0.003 |
| SOFA score on day 4 | 5 (3-8) | 6 (4-9) | <0.001 |
| Septic shock<sup>b</sup> | 203 (15.4) | 191 (19.5) | 0.01 |
| Prior ICU admission during hospital stay | 166 (12.6) | 118 (12.0) | 0.76 |
| **Therapeutic interventions** |  |  |  |
| High-dose corticosteroids<sup>b,c</sup> | 282 (21.3) | 278 (28.4) | <0.001 |
| Renal replacement therapy<sup>b</sup> | 156 (11.8) | 171 (17.4) | <0.001 |
| Mechanical ventilation on day 4 | 1030 (77.9) | 790 (80.6) | 0.13 |
| **Hematological parameters** |  |  |  |
| ANC [$\times 10^9/L$] on day 4 | 9.5 (6.9-1.2) | 8.3 (6.1-11.8) | <0.001 |
| AMC [ $\times 10^9/L$] on day 4 | 0.9 (0.7-1.2) | 0.6 (0.4-0.9) | <0.001 |
| Data are presented as medians (interquartile range) or absolute numbers (%). p values were calculated by nonparametric tests and Chi square tests, respectively. Abbreviations: ALC, absolute lymphocyte count; ICU, intensive care unit; APACHE, Acute Physiology and Chronic Health Evaluation; SOFA, sequential organ failure assessment; ANC, Absolute neutrophil count; AMC, Absolute monophil count.<br><sup>a</sup> use of immunosuppressive medication or systemic corticosteroids (prednisone >0.1 mg/kg for >3 months, prednisone >75 mg/day for >1 week, or equivalent), or chemotherapy and/or radiotherapy in the year before ICU admission<br><sup>b</sup> on one or more days in the course of the first 4 days in ICU<br><sup>c</sup> High dose corticosteroid was defined as a daily dose of hydrocortisone $\geq 200\text{mg}$ or equivalent |  |  |  |

Alluvial diagrams showed that lymphocyte counts remained relatively stable during the first four days in ICU for the majority of patients (figure 2), suggesting that ALC on day four was indeed a reasonable approximation of persistent lymphocytopenia as the exposure factor of interest. In fact, 790 (54.4%) patients having ALC <1.0 ×10^9//L upon ICU admission still had below-normal counts on day 4, whereas 659 (77.5%) patients having ALC ≥1.0 ×10^9//L on their first day remained in the normal range. Furthermore, among 1210 subjects for whom antecedent data were available, 344 (60.0%) individuals with low ALC in the week prior to ICU admission were still lymphocytopenic on day 4, whereas 446 (70.0%) individuals with higher ALC during this time period remained in the normal range throughout. Overall, the median duration of lymphocytopenia in the ICU (i.e., number of consecutive days with ALC <1.0 ×10^9/L) was 3 days (interquartile range 1-6) among the exposed patients. Onset of new lymphocytopenia occurring later than day 4 was rare; only 83 (7.1%) of all episodes of persistent lymphocytopenia met this criterion (see Supplementary Figure S1), whereas the daily proportion of patients in the ICU having lymphocytopenia remained relatively stable over time. Of the 980 patients with ALC <1.0 ×10^9/L on day 4, 461 (47%) had recovered to normal levels upon last measurement in the ICU. Descriptive analysis of persistent lymphocytopenia in relation to other hematological parameters did not reveal any clear relationship between level of ALC and absolute neutrophil or monocyte counts on day 4. Similarly, comparison of patients who later developed an ICU-AI to patients without ICU-AI did not reveal any clear patterns of neutrophil to lymphocyte ratio and monocyte to lymphocyte ratio distribution (Supplementary Figure S2).

**Figure 2.**
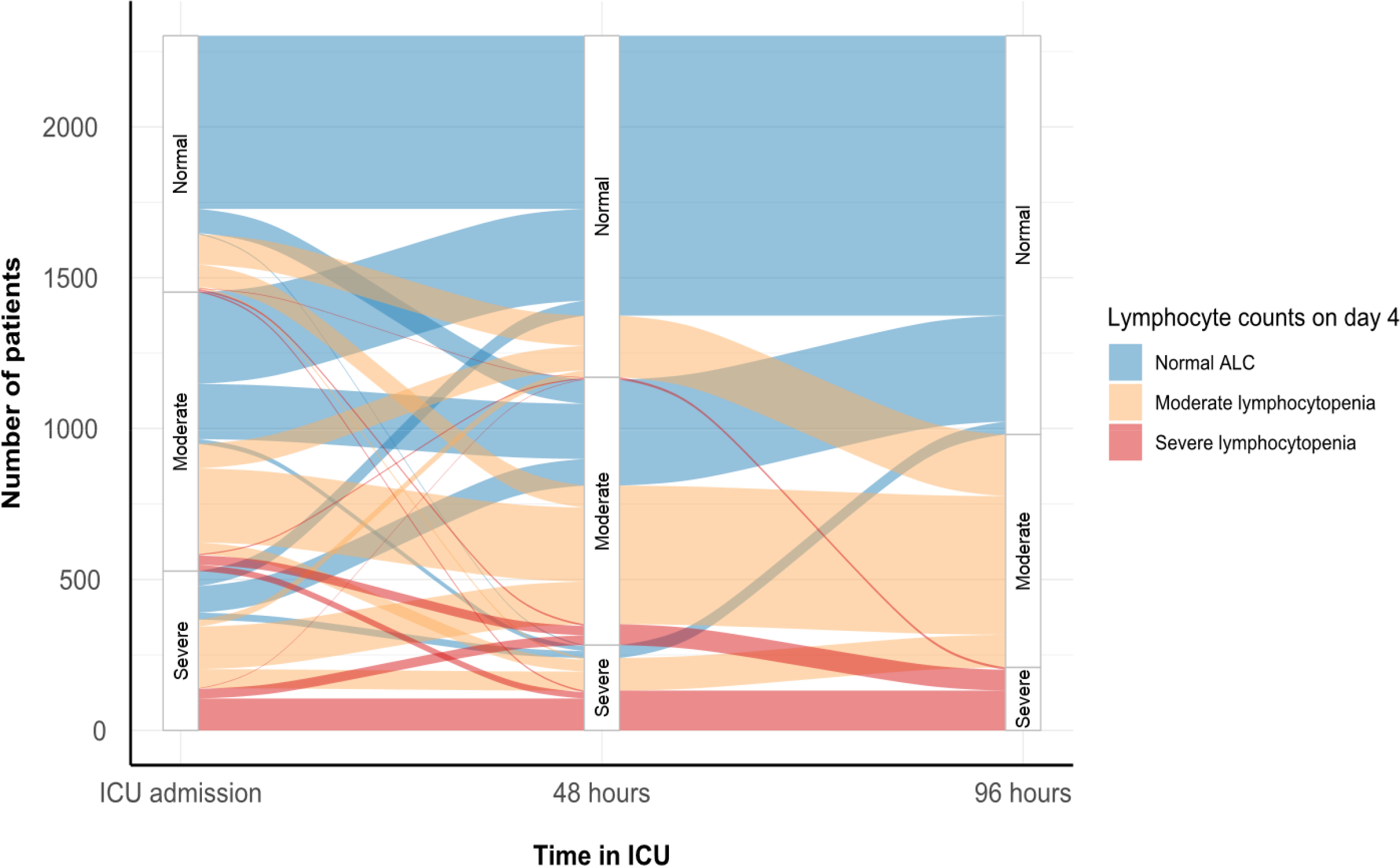
Evolution of lymphocyte counts in the first days of ICU admission. Alluvial plot illustrating the evolution of absolute lymphocyte counts (ALC) during the first 4 days in ICU. The thickness of the colored flow bars indicates the number of patients transitioning between each ALC level across time as indicated on the x-axis.

We observed a total of 597 episodes of ICU-AI amongst 453 (19.7%) patients, with 214 (21.8%) and 239 (18.0%) subjects with and without persistent lymphocytopenia being affected, respectively (p 0.03; table 2). The most common sources of infection included the respiratory tract (n=203), bloodstream (n=171), and abdomen (n=83). The median time to onset of (first) ICU-AI was similar in both groups. Crude mortality was 169 (17.2%) in patients with persistent lymphocytopenia compared to 167 (12.6%) in those without (p 0.003).

**Table 2.** ICU Outcomes

| **Table 2 ICU Outcomes** |  |  |  |
| --- | --- | --- | --- |
| ICU Outcomes | Normal ALC<br>N=1322 | Persistent<br>lymphocytopenia<br>N=980 | p |
| **Characteristics of ICU-acquired infections** |  |  |  |
| Patients with ICU-acquired infection | 239 (18.1) | 214 (21.8) | 0.03 |
| Day of first ICU-acquired infection | 8 (6 - 10) | 8 (5 -11) | 0.97 |
| Source of infection<sup>a</sup> |  |  |  |
| Pulmonary | 120 (38 %) | 83 (29 %) |  |
| ICU-acquired pneumonia | 102 (32 %) | 73 (26 %) |  |
| Bloodstream | 83 (26 %) | 88 (31 %) |  |
| Bacteremia | 13 (4 %) | 17 (6 %) |  |
| CRBSI | 70 (22 %) | 71 (25 %) |  |
| Abdominal | 34 (11 %) | 49 (17 %) |  |
| Neurological | 32 (10 %) | 13 (5 %) |  |
| Other<sup>b</sup> | 46 (15 %) | 49 (17 %) |  |
| **Other ICU-outcome** |  |  |  |
| Length of ICU stay in ICU survivors | 8 (5 – 13.5) | 9 (6 – 14.5) | 0.08 |
| ICU mortality | 167 (12.6) | 169 (17.2) | 0.002 |
| Data are presented as medians (interquartile range) or absolute numbers (%).ICU Intensive Care Unit, IQR Interquartile Range, HAP Hospital-acquired pneumonia, VAP Ventilator associated pneumonia, CRBSI catheter-related bloodstream infection. |  |  |  |
| <sup>a</sup> absolute number and percentage (%) of ICU-acquired infections |  |  |  |
| <sup>b</sup> Other infections include lung abscess, sinusitis, pharyngitis, tracheobronchitis, endocarditis, myocarditis, bone and joint infection, eye infection, reproductive tract infection, skin or soft tissue infections, postoperative wound infection, sepsis eci. |  |  |  |

Table 3 shows the results of the competing risk survival analyses. By cause-specific regression, persistent lymphocytopenia appeared to have no direct relation to daily risk of ICU-AI (crude CSHR 1.14, 95% CI, 0.95-1.37). However, crude analyses did reveal slight trends towards a lower daily probability of being discharged as well as an increased daily risk of dying in the ICU. Consequently, when these effects were combined, persistent lymphocytopenia was indeed crudely associated with a higher cumulative incidence of ICU-AI (SHR 1.23, 95% CI, 1.02-1.48, Supplementary Figure S2) However, after adjustment for potential confounders using multivariable models, all observed effects were considerably reduced and persistent lymphocytopenia no longer remained associated with an increased occurrence of ICU-AI, either directly (adjusted CSHR 1.08, 95% CI, 0.90-1.31) or indirectly (adjusted SHR 1.09, 95% CI, 0.90-1.32).

**Table 3.** Associations between lymphocytopenia and clinical outcome.

| **Table 3 Associations between lymphocytopenia and clinical outcome** |  |  |  |  |  |  |  |
| --- | --- | --- | --- | --- | --- | --- | --- |
|  | ICU-acquired infection |  | ICU mortality |  | ICU discharge |  | ICU-acquired infection |
|  | CSHR (95% CI) |  | CSHR (95% CI) |  | CSHR (95% CI) |  | SHR (95% CI) |
| Cox proportional model |  |  |  |  |  |  |  |
| Crude model | 1.14 | (0.95 - 1.37) | 1.32 | (1.00 - 1.73) | 0.85 | (0.77 - 0.94) | 1.23 (1.02 - 1.48) |
| Adjusted multivariable model <sup>a</sup> | 1.08 | (0.90 - 1.31) | 1.19 | (0.90 - 1.58) | 1.00 | (0.90 - 1.11) | 1.09 (0.91 - 1.32) |
| Data are presented as hazard ratios with 95% confidence intervals. The cause-specific hazard ratio estimates the direct effect of lymphocytopenia on clinical outcome (i.e., ICU-acquired infection, discharge alive or death). The subdistribution hazard ratio is a summary measure of the separate cause-specific hazards and estimates the overall risk of ICU-acquired infection in patients with lymphocytopenia while taking into account the competing events.<br>Abbreviations: CI, confidence interval; CSHR, cause-specific hazard ratio; ICU, intensive care unit; SHR, subdistribution hazard ratio<br><sup>a</sup>Baseline covariables adjusted for in the model were age, sex, Charlson Comorbidity Index, episode of sepsis before D4, immunosuppressant therapy prior to ICU admission, APACHE IV Acute Physiology Score, treatment with high-dose corticosteroids in first days of ICU admission, day 4 SOFA score |  |  |  |  |  |  |  |

We performed a number of sensitivity analyses to assess the robustness of these results. Neither use of different thresholds to define the exposure variable (i.e., persistent lymphocytopenia), nor restriction of the primary endpoint (i.e., ICU-AI) to include only specific types of infection changed our study findings (Supplementary Tables S1-6). Of note, a highly sensitive analysis that used a lymphocytopenia score to quantify duration and severity of lymphocytopenia also did not yield a positive association (Supplementary Table S7).

Finally, we performed a number of subgroup analyses to assess potential effect modification. The crude associations between persistent lymphocytopenia and the occurrence of ICU-AI (and death) were most pronounced for patients who had presented to the ICU with a primary admission diagnosis of sepsis, as compared to subgroups of patients having the highest disease severity scores or containing only those who had been admitted following trauma and/or major surgery (Supplementary Table S6). However, as for the primary analysis, none of these associations remained after adjustment for confounding.

## Discussion

We studied the prevalence, temporal evolution and clinical correlates of persistent lymphocytopenia during critically illness and assessed its potential etiological role in the development of subsequent ICU-AI. Although our analyses revealed a crude association between the occurrence of persistent lymphocytopenia and the cumulative incidence of nosocomial infections (and death) in the ICU, perturbations in ALC do not appear to constitute a direct risk factor for infection after adjustment for competing risks and confounders.

The prevalence of lymphocytopenia (ALC <1.0 ×10^9/L) was high in our cohort (with a peak prevalence of 63.1% at ICU admission), and 79.7% of patients were affected on at least a single day during their stay in the ICU. This is in line with previous research conducted in various populations of critically ill patients, with reported prevalences varying between 52% and 63% [4, 16, 20, 26]. Our observations also suggest that lymphocytopenia generally develops early in the course of critical illness, and thus most likely occurs in response to the primary insult that led to ICU admission. This supports the notion that induction of many host responses in critical illness may occur simultaneously [1, 27] rather than successively as conventionally presumed [28]. When severity of lymphocytopenia was compared between patients with infectious, surgical or traumatic reasons for admission, we did not observe any significant differences. If differences in host response in the presence of critical illness indeed exists between various ICU etiologies, they are likely to lie deeper than the ALC. Interestingly, although not statistically significant, post-hoc subgroup analysis yielded prominent trends in the direct effect between exposure and ICU mortality in patients admitted with sepsis. As no association was found between persistent lymphocytopenia and ICU-AI, the possible association between persistent lymphocytopenia and ICU mortality does not appear to be facilitated through the development of new infectious complications in the ICU. Alternative mechanisms related to lymphocytopenia, such as impaired ability to clear the initial infection upon admission, could, however, play a role. Indeed, a postmortem study in sepsis non-survivors revealed that 80% of study patients still had a septic focus on autopsy [29]. Although the role of lymphocytopenia and failure to eliminate infection was not described in this study, there is evidence that indicates defects in lymphocyte populations can lead to deficient bacterial clearance in sepsis [30-32].

Our results argue against a mechanistic link between persistent lymphocytopenia and ICU-AI and contest the rationale behind recently proposed immune-adjuvant therapies aiming to modulate critical illness-related immunosuppression and boost lymphocyte function [15, 33]. A recent phase IIb randomized control trial showed that 4 weeks of recombinant human interleukin-7 (IL-7) therapy in patients with septic shock and severe lymphocytopenia is safe and may restore ALC [34]. However, no effect on the incidence of subsequent hospital-acquired infections —a secondary aim of the study— could be demonstrated. Two other forms of antibody-mediated immunotherapy blocking the programmed cell death-1 (PD-1) receptor and its ligand (PD-L1) with the goal of preventing lymphocyte exhaustion, were also well-tolerated, but failed to induce changes in ALC [35, 36]. Although these studies were limited by small sample sizes and the true effectiveness of both IL-7 and anti-PD-1/PD-L1 remains to be determined in larger randomized control trials, the results from the first clinical trials of interventions that aimed to augment ALC appear to agree with our findings and imply that ALC alone may not be the ideal therapeutic target to reduce nosocomial infections and mortality in the ICU. However, a possible beneficial effect arising from the prevention of later nosocomial infections (occurring after ICU discharge) cannot be excluded based on our findings.

Although ALC is clearly not associated with ICU-AI, other indices of lymphocyte function could provide insight into the possible role of critical illness-related immunosuppression and adverse outcomes in the ICU. As full differential WBC analysis is not routinely performed in our hospital, we could not investigate perturbations in different subpopulations of lymphocytes, such as regulatory CD4 T-cells or memory B-cells. Whereas ALC only measures the absolute number of circulating lymphocytes, the relative abundance or absence of certain lymphocyte subtypes or the expression of specific phenotypes may reveal more about the functionality of the immune system and may act as prognostic markers of nosocomial infection and mortality. [8-10, 33, 37-39] While these biochemical markers may not be feasible candidates for use in everyday clinical practice, they could be utilized as part of prognostic and predictive enrichment strategies in clinical trials and precision medicine.

A few limitations inherent to our study population and design should be noted. First, the study was conducted in a single center, where selective decontamination of the digestive tract (SDD) was routinely applied during ICU admission. SDD reduces (1) colonization of the respiratory and intestinal tract with potentially pathogenic microorganisms, (2) incidence of ICU-acquired bacteremia, and (3) mortality in ICU settings with a low prevalence of multidrug-resistant bacteria [40], thereby limiting the generalizability of our results. Second, we did not collect data beyond ICU discharge, and thus cannot rule out potential associations between lymphocytopenia and the occurrence of late-onset nosocomial infections. Third, we did not explicitly model the dynamic relation between timing of onset and/or duration of lymphocytopenia and ICU-AI occurrence. However, even though we observed some fluctuations in day-to-day lymphocyte counts, by using ALC on day 4 as a pragmatic exposure definition we captured >90% of all episodes of persistent lymphocytopenia that ever occurred in the ICU (figure S1). We therefore believe a more complex analytical approach (such as a multistate model), which would require several bold *a priori* assumptions about the hypothetical temporal relation between exposure and outcome, is not only unfeasible but also unlikely to change our findings. Finally, we also cannot exclude the possibility of residual confounding in our multivariable analyses, which could have led to an underestimation of true effect.

To our knowledge, our study is the largest cohort to study the association of lymphocytopenia and ICU-AI in a general ICU population. Daily measurement of ALC enabled us to investigate the time course of lymphocytopenia and provide face validity to our day-4 exposure definition, as well as perform extensive sensitivity analyses using alternative criteria. Further strengths of our study include the use of methodologies to minimize bias by confounding and/or competing risks. Moreover, all episodes of ICU-AI were prospectively registered and carefully adjudicated by trained observers, which reduces the risk of misclassification of the outcome.

## Conclusion

Persistent lymphocytopenia occurs commonly in critically ill patients during their first days in ICU, but does not appear to play an important role in the development of ICU-acquired infections.

## Data Availability

As this study was carried out on the basis of analysis of routinely collected data by the usual care team, consent for sharing of anonymized patient level data is not available.

## ABBREVIATIONS

ALC: Absolute lymphocyte counts
APACHE: Acute physiology and chronic health evaluation
CSHR: Cause-specific hazard ratio
CI: Confidence interval
ICU: Intensive care unit
ICU-AI: Intensive care unit-acquired infections
IL-7: Interleukin-7
IQR: Interquartile range
MARS: Molecular Diagnosis and Risk Stratification of Sepsis cohort
PD-1: Programmed cell death-1 receptor
PD-L1: Programmed cell death-1 receptor ligand
SDD: Selective decontamination of the digestive tract
SHR: Subdistribution hazard ratio
SOFA: Sequential organ failure assessment

## REFERENCES

1. Xiao W, Mindrinos MN, Seok J, et al: Inflammation, Host Response to Injury Large-Scale Collaborative Research P. A genomic storm in critically injured humans. J Exp Med 2011; 208:2581–2590. doi:10.1084/jem.20111354.

2. Otto GPS, Sossdorf M, Claus RA, et al: The late phase of sepsis is characterized by an increased microbiological burden and death rate. Crit Care 2011; 15:R183. doi:10.1186/cc10332.

3. Stortz JA, Murphy TJ, Raymond SL, et al: Evidence for Persistent Immune Suppression in Patients Who Develop Chronic Critical Illness After Sepsis. Shock 2018; 49:249–258. doi:10.1097/SHK.0000000000000981.

4. Adrie C, Lugosi M, Sonneville R, et al: Persistent lymphopenia is a risk factor for ICU-acquired infections and for death in ICU patients with sustained hypotension at admission. Ann Intensive Care 2017; 7:30. doi:10.1186/s13613-017-0242-0.

5. Vincent JL, Rello J, Marshall J, et al: International Study of the Prevalence and Outcomes of Infection in Intensive Care Units. JAMA 2009; 302:2323–2329. doi:10.1001/jama.2009.1754.

6. Hotchkiss RS, Swanson PE, Freeman BD, et al: Apoptotic cell death in patients with sepsis, shock, and multiple organ dysfunction. Crit Care Med 1999; 27:1230–51

7. Hotchkiss RS, Tinsley KW, Swanson PE, et al: Sepsis-induced apoptosis causes progressive profound depletion of B and CD4+ T lymphocytes in humans. J Immunol 2001; 166:6952–63

8. Guignant C, Lepape A, Huang X, et al: Programmed death-1 levels correlate with increased mortality, nosocomial infection and immune dysfunctions in septic shock patients. Crit Care 2011; 15:R99. doi:10.1186/cc10112.

9. Frattari A, Polilli E, Primiterra V, et al: Analysis of peripheral blood lymphocyte subsets in critical patients at ICU admission: A preliminary investigation of their role in the prediction of sepsis during ICU stay. Int J Immunopathol Pharmacol 2018; 32:2058738418792310. doi:10.1177/2058738418792310.

10. Gouel-Cheron A, Venet F, Allaouchiche B, et al: CD4+ T-lymphocyte alterations in trauma patients. Crit Care 2012; 16:432. doi:10.1186/cc11376.

11. Cheadle WG, Pemberton M, Robinson D, et al: Lymphocyte subset responses to trauma and sepsis. J Trauma 1993; 35:844–849

12. van den Broek T, Borghans JAM, van Wijk F: The full spectrum of human naive T cells. Nat Rev Immunol 2018; 18:363–373. doi:10.1038/s41577-018-0001-y.

13. Wherry EJ, Kurachi M: Molecular and cellular insights into T cell exhaustion. Nat Rev Immunol 2015; 15:486-499. doi:10.1038/nri3862.

14. Williams MA, Bevan MJ: Effector and memory CTL differentiation. Annu Rev Immunol 2007; 25:171–192.

15. Patil NK, Bohannon JK, Sherwood ER: Immunotherapy: A promising approach to reverse sepsis-induced immunosuppression. Pharmacol Res 2016; 111:688-702. doi:10.1016/j.phrs.2016.07.019.

16. Drewry AM, Samra N, Skrupky LP, et al: Persistent lymphopenia after diagnosis of sepsis predicts mortality. Shock 2014; 42:383-391. doi:10.1097/SHK.0000000000000234.

17. Felmet KA, Hall MW, Clark RS, et al: Prolonged lymphopenia, lymphoid depletion, and hypoprolactinemia in children with nosocomial sepsis and multiple organ failure. J Immunol 2005; 174:3765–3772

18. Heffernan DS, Monaghan SF, Thakkar RK, et al: Failure to normalize lymphopenia following trauma is associated with increased mortality, independent of the leukocytosis pattern. Crit Care 2012; 16:R12. doi:10.1186/cc11157.

19. Vulliamy PE, Perkins Z, Brohi K, et al: Persistent lymphopenia is an independent predictor of mortality in critically ill emergency general surgical patients. Eur J Trauma Emerg Surg 2016; 42:755–60.

20. Bermejo-Martin JF, Cilloniz C, et al: Lymphopenic Community Acquired Pneumonia (L-CAP), an Immunological Phenotype Associated with Higher Risk of Mortality. EBioMedicine 2017; 24:231–236. doi:10.1016/j.ebiom.2017.09.023.

21. Chung KP, Chang HT, Lo SC, et al: Severe lymphopenia is associated with elevated plasma interleukin-15 levels and increased mortality during severe sepsis. Shock 2015; 43:569–575. doi:10.1097/SHK.0000000000000347.

22. ten Berg MJ, Huisman A, van den Bemt PM, et al: Linking laboratory and medication data: new opportunities for pharmacoepidemiological research. Clin Chem Lab Med 2007; 45:13–9.

23. Vasu S, Caligiuri MA. Lymphocytosis and Lymphocytopenia. In: Kauhansky K, Lichtman MA, Prchal JT, Levi MM, Press OW, Burns LJ, Caligiuri M, editors. williams Hematology, 9^th^ ed. New York, NY:McGraw-Hill Education; 2015. Available from: https://accessmedicine.mhmedical.com/content.aspx?bookid=1581&sectionid=108068723#1121097296. Accessed April 6, 2020.

24. Klein Klouwenberg PM, Ong DS, Bos LD, et al: Interobserver agreement of Centers for Disease Control and Prevention criteria for classifying infections in critically ill patients. Crit Care Med 2013; 41:2373–2378. doi:10.1097/CCM.0b013e3182923712.

25. Inoue S, Suzuki-Utsunomiya K, Okada Y, et al: Reduction of immunocompetent T cells followed by prolonged lymphopenia in severe sepsis in the elderly. Crit Care Med 2013; 41:810–819. doi:10.1097/CCM.0b013e318274645f.

26. de Jager Cpc, van Wijk Ptl, Mathoera RB, et al: Lymphocytopenia and neutrophil-lymphocyte count ratio predict bacteremia better than conventional infection markers in an emergency care unit. Crit Care 2010; 14:R192. doi:10.1186/cc9309.

27. Frencken JF, van Vught LA, Peelen LM, et al: An Unbalanced Inflammatory Cytokine Response Is Not Associated With Mortality Following Sepsis: A Prospective Cohort Study. Crit Care Med 2017; 45:e493–e499. doi:10.1097/CCM.0000000000002292.

28. Hotchkiss RS, Monneret G, Payen D. Immunosuppression in sepsis: a novel understanding of the disorder and a new therapeutic approach. The Lancet Infectious Diseases 2013; 13:260–268. doi:10.1016/S1473-3099(13)70001-X.

29. Torgersen C, Moser P, Luckner G, et al: Macroscopic postmortem findings in 235 surgical intensive care patients with sepsis. Anesth Analg 2009; 108:1841–1847. doi:10.1213/ane.0b013e318195e11d.

30. Kelly-Scumpia KM, Scumpia PO, Weinstein JS, et al: B cells enhance early innate immune responses during bacterial sepsis. J Exp Med 2011; 208:1673–1682. doi:10.1084/jem.20101715.

31. Rauch PJ, Chudnovskiy ., Robbins CS, et al: Innate Response Activator B Cells Protect Against Microbial Sepsis. Science 2012; 335:597–601. doi:10.1126/science.1215173.

32. Wellmer A, von Mering M, Spreer A, et al: Experimental pneumococcal meningitis: impaired clearance of bacteria from the blood due to increased apoptosis in the spleen in Bcl-2-deficient mice. Infect Immun 2004; 72:3113–3119

33. Shankar-Hari M, Datta D, Wilson J, et al: Early PREdiction of sepsis using leukocyte surface biomarkers: the ExPRES-sepsis cohort study. Intensive Care Med 2018; 44:1836–1848. doi:10.1007/s00134-018-5389-0.

34. Francois B, Jeannet R, Daix T, et al: Interleukin-7 restores lymphocytes in septic shock: the IRIS-7 randomized clinical trial. JCI Insight 2018; 3. doi:10.1172/jci.insight.98960.

35. Hotchkiss RS, Colston E, Yende S, et al: Immune checkpoint inhibition in sepsis: a Phase 1b randomized study to evaluate the safety, tolerability, pharmacokinetics, and pharmacodynamics of nivolumab. Intensive Care Med 2019; 45:1360–1371. doi:10.1007/s00134-019-05704-z.

36. Hotchkiss RS, Colston E, Yende S, et al: Immune Checkpoint Inhibition in Sepsis: A Phase 1b Randomized, Placebo-Controlled, Single Ascending Dose Study of Antiprogrammed Cell Death-Ligand 1 Antibody (BMS-936559). Crit Care Med 2019; 47:632–642. doi:10.1097/CCM.0000000000003685.

37. Boomer JS, Shuherk-Shaffer J, Hotchkiss RS, et al: A prospective analysis of lymphocyte phenotype and function over the course of acute sepsis. Crit Care 2012; 16:R112. doi:10.1186/cc11404.

38. Conway Morris A, Datta D, Shankar-Hari M, et al: Cell-surface signatures of immune dysfunction risk-stratify critically ill patients: INFECT study. Intensive Care Med 2018; 44:627–635. doi:10.1007/s00134-018-5247-0.

39. Dong X, Liu Q, Zheng Q, et al: Alterations of B Cells in Immunosuppressive Phase of Septic Shock Patients. Crit Care Med. 2020; 48:815-821. doi:10.1097/CCM.0000000000004309.

40. Wittekamp BHJ, Oostdijk EAN, Cuthbertson BH, et al: Selective decontamination of the digestive tract (SDD) in critically ill patients: a narrative review. Intensive Care Med 2019; 46:343–349. doi:10.1007/s00134-019-05883-9.

